# Automated classification of neurodegenerative parkinsonian syndromes using multimodal magnetic resonance imaging in a clinical setting

**DOI:** 10.1101/2020.03.27.20042671

**Authors:** Lydia Chougar, Johann Faouzi, Nadya Pyatigorskaya, Rahul Gaurav, Emma Biondetti, Marie Villotte, Romain Valabrègue, Jean-Christophe Corvol, Alexis Brice, Louise-Laure Mariani, Florence Cormier, Marie Vidailhet, Gwendoline Dupont, Ines Piot, David Grabli, Christine Payan, Olivier Colliot, Bertrand Degos, Stéphane Lehéricy

## Abstract

**Background:** Several studies have shown that machine learning algorithms using MRI data can accurately discriminate parkinsonian syndromes. Validation under clinical conditions is missing.

**Objectives:** To evaluate the accuracy for the categorization of parkinsonian syndromes of a machine learning algorithm trained with a research cohort and tested on an independent clinical replication cohort.

**Methods:** 361 subjects, including 94 healthy controls, 139 patients with PD, 60 with PSP with Richardson’s syndrome, 41 with MSA of the parkinsonian variant (MSA-P) and 27 with MSA of the cerebellar variant (MSA-P), were recruited. They were divided into a training cohort (n=179) scanned in a research environment, and a replication cohort (n=182), scanned in clinical conditions on different MRI systems. Volumes and DTI metrics in 13 brain regions were used as input for a supervised machine learning algorithm.

**Results:** High accuracy was achieved using volumetry in the classification of PD versus PSP, PD versus MSA-P, PD versus MSA-C, PD versus atypical parkinsonian syndromes and PSP versus MSA-C in both cohorts, although slightly lower in the replication cohort (balanced accuracy: 0.800 to 0.915 in the training cohort; 0.741 to 0.928 in the replication cohort). Performance was lower in the classification of PSP versus MSA-P and MSA-P versus MSA-C. When adding DTI metrics, the performance tended to increase in the training cohort, but not in the replication cohort.

**Conclusions:** A machine learning approach based on volumetric and DTI data can accurately classify subjects with early-stage parkinsonism, scanned on different MRI systems, in the setting of their clinical workup.

## Introduction

The diagnosis of idiopathic Parkinson’s disease (PD) and atypical parkinsonism, whose most frequent types are progressive supranuclear palsy (PSP) and multiple system atrophy (MSA), rely on clinical criteria.^1–3^ At initial presentation, a correct diagnosis may be difficult with high clinical uncertainty. However, an accurate diagnosis is crucial to provide prognostic assessment and adequate counseling and for the categorization of patients before inclusion in therapeutic trials. It has been shown that the diagnostic accuracy of PD was much improved when clinical experts in movement disorders made the diagnosis with a sensitivity of 91.1% and a specificity of 98.4%^1,4^

Degeneration of dopaminergic neurons within the substantia nigra *pars compacta* is the hallmark of all neurodegenerative parkinsonian syndromes.^5–7^ While PD patients only exhibit nigral abnormalities in a limited number of small brainstem nuclei at the early stages of the disease,^5–8^ PSP patients show a larger involvement of other structures, predominating in the midbrain, the dentate nucleus and superior cerebellar peduncles ^6,7,9–11^ and MSA patients are characterized by damages particularly affecting the posterior putamen in the parkinsonian variant (MSA-P) and the pons, middle cerebellar peduncles and cerebellum in the cerebellar variant (MSA-C).^6,7,12^

Multimodal magnetic resonance imaging (MRI) can detect these differential patterns of brain involvement.^13–17^ *In vivo* neuroimaging biomarkers that correlate with neuropathological alterations can be extracted from MRI data sensitive to several properties of the underlying brain tissue. Indeed, brain atrophy is visible on T1-weighted images, tissue microstructure alterations are detected using diffusion-weighted images, and iron deposition can be evidenced using iron-sensitive sequences.^13–17^

Recently, several studies have demonstrated that machine learning algorithms trained with MRI data could differentiate between parkinsonian syndromes with high accuracy.^18–25^ Nevertheless, most of these studies were based on one single type of MRI acquisition protocol, either T1-weighted volumetry^18–20^ or diffusion-weighted data,^25^ with three studies using a multimodal approach combining volumetry and diffusion^21^ or including R2* relaxometry^22^ or spectroscopy.^23^ Two studies have relied on large cohorts, including 1002^25^ or 464 subjects,^19^ whereas others studies have investigated smaller samples.^18,20–23^ Only two studies have included at the same time PD, PSP, MSA-P and MSA-C subjects,^19,23^ while other studies have not differentiated between MSA-P and MSA-C, ^18^ have only included MSA-P patients ^25^ or MSA-C patients ^24^ or have not included MSA ^20,21^ or PSP patients. ^22^ Moreover, these studies have mainly been designed in a research environment and tested without an independent replication cohort.^19–22^ To translate an automated machine-learning-based classification into clinical practice, there is a need for multimodal studies based on large cohorts and tested on clinical populations including all types of parkinsonian syndromes.

Our objective was to evaluate the predictive performance of a machine learning algorithm for the categorization of parkinsonian syndromes including MSA-C, MSA-P and PSP compared with PD and healthy controls (HC). This algorithm was trained on a research cohort and tested on an independent replication cohort scanned on different MRI scanners in clinical conditions in a Neuroradiology department, using volumetric and diffusion-weighted MRI data.

## Material and methods

### Population

Two populations of subjects were included: a training cohort to train and validate the algorithm and a replication cohort to independently evaluate the algorithm’s performance. Subjects in the training cohort were enrolled in the frame of three research studies conducted between 2007 and 2012 at the Brain and Spine Institute (ICM), Paris: Genepark (LSHB-CT-2006-037544), BBBIPPS (DGS 2006/0524) and Nucleipark (RCB 2009-A00922-55). Inclusion criteria for patients were: 1) a diagnosis of PD, PSP or MSA established by movement disorders specialists according to published consensus criteria for PD (UK Parkinson’s Disease Society Brain Bank clinical diagnostic criteria ^1^ and no or minimal cognitive disturbances with Mini Mental State Examination>24), PSP with a Richardson syndrome (National Institute of Neurological Disorders and Stroke criteria) ^26^ or MSA (Second consensus statement on the diagnosis of multiple system atrophy).^3^

Subjects in the replication cohort were consecutively and prospectively enrolled between 2013 and 2019 in the movement disorders clinic and the Neuroradiology department of the Pitié-Salpêtrière University Hospital, Paris. Diagnosis of probable PD, PSP or MSA were retrospectively established in 2019 by movement disorders specialists according to the above mentioned clinical criteria based on all available clinical data throughout the follow-up.

The clinical examination included the Unified Parkinson’s Disease Rating Scale part III scores (UPDRS III). Baseline MRI scans were obtained on the same day of the clinical examination. For both cohorts, HCs matched for age with patients were included. They had no history of neurological or psychiatric disease. Subjects were excluded if they had any additional neurological disorder including stroke or brain tumor on MRI examinations.

Institutional review boards approved the studies (Genepark: CPP Paris II, 2007-A00208-45; BBBIPPS: CPP Paris VI, P040410 – 65-06; Nucleipark: CPP Paris VI, 65-09; Park Atypique: CPP Ile-de-France VI, 08012015). Written informed consent was obtained from all participants.

### Image acquisition

Subjects in the training cohort were scanned at the Brain and Spine Institute using a 3.0T TRIO (Siemens, Erlangen, Germany) with a 32-channel head coil. Subjects in the replication cohort were scanned in clinical conditions for diagnostic purposes at the hospital’s Neuroradiology department using four MRI systems: 1) 1.5T GE OPTIMA 450 (General Electrics Medical Systems, Milwaukee, USA) using a 32-channel head coil, 2) 3T GE SIGNA HDxt (General Electrics Medical Systems, Milwaukee, USA) using a 32-channel head coil, 3) 3T GE Discovery MR750 (PET/MR) (General Electrics Medical Systems, Milwaukee, USA), using a 8-channel head coil, 4) 3T Siemens SKYRA (Siemens, Erlangen, Germany), using a 64-channel head coil. HCs in the replication cohort were scanned twice during different sessions on 3T SKYRA and 3T SIGNA MRI systems.

All subjects were scanned using a standardized protocol adapted to each scanner. The protocols included a high-resolution T1-weighted gradient-recalled echo sequence (Magnetization-prepared rapid acquisition with gradient-recalled echo, MPRAGE, or Spoiled Gradient Recalled Acquisition in Steady State, SPGR) and diffusion tensor imaging (DTI) with 15 (1.5T GE), 30 (3T SKYRA, 3T GE SIGNA and GE PET/MR system), 60 or 64 (3T TRIO) gradient-encoding directions. The acquisition parameters are provided in the supplementary Table S1. DTI sequences were missing in 19 patients in the training cohort and 17 patients in the replication cohort. Quality control was performed by visual inspection; T1-weighted and diffusion-weighted images with significant motion artifacts or image distortions were excluded.

### Data processing and analysis

Image processing and analysis were performed using in-house software written in Matlab (R2017b, The MathWorks, Inc., Natick, MA, USA).

T1-weighted images were automatically segmented using Freesurfer (http://freesurfer.net/, MGH, Boston, MA, USA). DTI preprocessing was performed using the FMRIB Software Library (FSL) v5.0 (FMRIB, Oxford, UK). Motion and eddy currents were corrected using the eddycor function. Fractional anisotropy (FA) and diffusivity maps were computed with the DTIfit function for the entire brain volume. The diffusion maps were coregistered to the 3D T1-weighted volume using the SPM coregister function. The volumes, mean values of FA, mean diffusivity (MD), axial diffusivity (AD) and radial diffusivity (RD) were calculated in all segmented regions of interest (ROI).

Thirteen ROI known for being involved in parkinsonian syndromes were included: midbrain, pons, third ventricle (V3), fourth ventricle (V4), superior cerebellar peduncles (SCP), cerebellum white matter (including the middle cerebellar peduncles), putamen, posterior putamen, caudate, thalamus, pallidum, precentral cortex and insular cortex.

### SVM classification

Using the scikit-learn package,^27^ four supervised machine learning algorithms were developed: a logistic regression, a support-vector machine (SVM) with a linear kernel, a SVM with a radial basis function kernel and a random forest. They were trained and validated on the training cohort, then tested on the replication cohort. Only the results obtained using the SVM with a linear kernel were reported as it tended to provide the best performance (results with the three other algorithms provided in the supplementary Tables S2 and S3).

The cross-validation procedure on the training cohort included two nested loops: an outer loop evaluating the classification performances and an inner loop used to optimize the hyperparameters of the algorithms. More precisely, repeated stratified random splits with five repetitions were used for outer cross-validation. For hyperparameter optimization, an inner loop with five-fold cross-validation was used. For each split, the model allowing the highest area under the ROC curve (AUC) among the five selected models was selected.

The input features were volumes, including the ratio between midbrain and pons volumes, FA, MD, AD and RD in the different ROI. Age and sex were added as covariables in the algorithm.

All subjects volumetric and DTI values (subjects referring to patients and controls) in the replication cohort were normalized by subjects values in the training cohort (subtraction of each individual value by the mean of all subjects in the training cohort, divided by the standard deviation).

To harmonize data across scanners and reduce potential scanner-dependent effects in the images, a separate analysis was conducted on a subset of the replication cohort including subjects scanned on the machines where HCs were available, namely SKYRA (n=84) and SIGNA (n=79). First, subject values in the replication cohort were normalized by subject values in the training cohort. Second, a normalization of subjects data by HCs data in both cohorts was performed (subtraction between each patient’s value and the mean of HCs, divided by the standard deviation of HCs).

The following classifications were evaluated: PD versus PSP, PD versus MSA-P, PD versus MSA-C, PD versus atypical parkinsonism, PSP versus MSA-P, PSP versus MSA-C and MSA-P versus MSA-C.

### Weighting factors

Weighting factors for each feature were extracted from SVM with linear kernel training. They reflected the contribution of each feature to group differentiation. A rescaling to a range of −1 to +1 was done to highlight the relative importance of each feature: the higher the absolute value, the bigger the contribution of the feature. This assertion relied on the assumption that each feature had the same scale, which was a reasonable assumption because of the standardization of each feature. When the coefficient was positive, the algorithm favored the first group if the value of the feature was high or the second group if the value was low. Conversely, when the coefficient was negative, the algorithm favored the second group if the value of the feature was high, or the first group if the value was low.^19^

### Statistical analyses

To evaluate the performance of the algorithm, Receiver operating characteristic (ROC) curves were generated; balanced accuracy (BA), AUC, sensitivity and specificity were calculated for each group comparison. The BA was defined as the average sensitivity and specificity in each group. BA avoids overestimation of classification performance due to imbalanced group sizes.^19^

## Results

### Clinical characteristics of the patients

In total, 361 subjects were analyzed, divided into a training cohort (n=179) with 72 HCs, 63 patients with PD, 21 with PSP, 11 MSA-P and 12 MSA-C, and a replication cohort (n=182) with 22 HCs, 76 patients with PD, 39 with PSP, 30 MSA-P and 15 MSA-C (Table 1).

**Table 1:** Demographic and clinical characteristics of the population.

|  | Training cohort |  |  |  |  | Replication cohort |  |  |  |  |
| --- | --- | --- | --- | --- | --- | --- | --- | --- | --- | --- |
| Groups | HC | PD | PSP | MSA-P | MSA-C | HC | PD | PSP | MSA-P | MSA-C |
| n | 72 | 63 | 21 | 11 | 12 | 22 | 76 | 39 | 30 | 15 |
| Age at scan | 60.8 ± 8.2 <sup>a</sup> | 60.7 ± 9.7 <sup>a</sup> | 65.6 ± 9.1 <sup>a</sup> | 62.8 ± 7.1 | 60.3 ± 7.4 | 64.7 ± 7.3 | 66.6 ± 10.8 | 71.4 ± 5.9 | 63.1 ± 7.4 | 60.6 ± 9.1 |
| Sex ratio (M/F) | 0.5 (24/48) | 1.4 (37/26) <sup>b</sup> | 0.9 (10/11) | 0.6 (4/7) | 1.4 (7/5) | 1.2 (12/10) | 1.7 (48/28) | 4.6 (32/7) <sup>c</sup> | 3.3 (23/7) | 2.0 (10/5) |
| Disease duration |  | 6.0 ± 4.0 <sup>d</sup> | 4.2 ± 1.9 | 5.0 ± 1.9 <sup>d</sup> | 5.2 ± 1.8 <sup>d</sup> |  | 4.3 ± 3.4 | 3.4 ± 1.8 | 3.5 ± 1.7 | 2.6 ± 1.6 |
| UPDRS III scores | 0.4 ± 0.7 | 21.0 ± 13.1 <sup>e</sup> | 38.5 ± 15.7 | 42.2 ± 13.9 | 47.4 ± 14.4 | 0.1 ± 0.3 | 22.0 ± 10.2 | 32.2 ± 16.9 <sup>f</sup> | 24.6 ± 10.4 | 16.7 ± 7.7 |

Age at MRI scan, disease duration and UPDRS III scores are expressed in mean ± standard deviation. They were compared using one way linear analysis of variance (ANOVA), followed by post hoc Student tests if case of significance. Sex were compared using a Chi-squared test.
<sup>a</sup> p<0.05 for HCs, PD and PSP patients between both cohorts
<sup>b</sup> p<0.001 for HCs versus PD in the training cohort
<sup>c</sup> p<0.05 for HCs versus PSP in the replication cohort
<sup>d</sup> p<0.05 for PD, MSA-P and MSA-C patients between both cohorts
<sup>e</sup> $p < 0.001$ for PD versus PSP, MSA-P and MSA-C in the training cohort
<sup>f</sup> $p = .01$ for PSP versus MSA-C in the replication cohort
HC: healthy controls; PD: Parkinson's disease; PSP: Progressive supranuclear palsy; MSA-C: cerebellar variant of multiple system atrophy; MSA-P: parkinsonian variant of multiple system atrophy; UPDRS III: Unified Parkinson's Disease Rating Scale part III; M: male; F: female.

In both cohort, there was no significant difference between patient groups in terms of age, sex and disease duration. In the training cohort, PD patients had lower UPDRS scores (21.0 ± 13.1) compared to PSP (38.5 ± 15.7, p<0.001), MSA-P (42.2 ± 13.9, p<0.001) and MSA-C (47.4 ± 14.4, p<0.001) patients. In the replication cohort, MSA-C patients had lower UPDRS scores than PSP patients (16.7 ± 7.7 versus 32.2 ± 16.9, p=0.0014). No other difference was seen in both cohorts

When comparing disease groups between both cohorts, HCs (64.7 ± 7.3 versus 60.8 ± 8.2, p=0.008), PD (66.6 ± 10.8 versus 60.7 ± 9.7, p=0.001) and PSP patients (71.4 ± 5.9 versus 65.6 ± 9.1, p=0.013) were older in the replication cohort. The disease durations were shorter for PD (4.3 ± 3.4 versus 6.0 ± 4.0, p=0.014), MSA-P (3.5 ± 1.7 versus 5.0 ± 1.9, p=0.004) and MSA-C (2.6 ± 1.6 versus 5.2 ± 1.8, p=0.012) patients in the replication cohort compared to the training cohort. PSP patients also had a shorter disease duration in the replication cohort though the difference was not significant (3.4 ± 1.8 versus 4.2 ± 1.9, p=0.088). UPDRS scores were lower in the replication cohort for MSA-P (24.6 ± 10.4 versus 42.2 ± 13.9, p<0.001) and MSA-C (16.7 ± 7.7 versus 47.4 ± 14.4, p=0.003) patients.

### Performances in the training cohort and the whole replication cohort

In the training cohort, when using volumetric data solely, the BAs ranged between 0.800 and 0.906, with AUCs between 0.945 and 1.000. The best classification performances were obtained in decreasing order for PD versus MSA-C (BA: 0.915; AUC: 1.000), PD versus PSP (BA: 0.907; AUC: 0.981), PSP versus MSA-C (BA: 0.850; AUC: 0.983), PD versus atypical parkinsonian syndromes (BA: 0.826; AUC: 0.945) and PD versus MSA-P (BA: 0.800; AUC: 0.946). Including DTI metrics improved BAs, the best differentiation being achieved for the classifications of PD versus PSP (BA: 0.935; AUC: 0.973) and PD versus MSA-C (BA: 0.923, AUC: 1.000) (Table 2). For the classifications of PSP versus MSA-P and MSA-C versus MSA-P, the performances were lower, with BAs equal to 0.660 and 0.733 and AUCs equal to 0.640 and 0.833, respectively. Combining volumetric and diffusion-weighted data did not improve BAs (Table 2).

**Table 2:** Performances of the linear SVM classification in the training cohort and in the whole replication cohort using volumetric data and the combination of volumetric and DTI data.

| **Disease groups** | **Volumetry only** |  | **Volumetry & DTI** |  |
| --- | --- | --- | --- | --- |
| **PD vs PSP** | **Training** | **Test** | **Training** | **Test** |
| **BA** | 0.906 (0.053) | 0.857 | 0.935 (0.057) | 0.673 |
| **AUC** | 0.981 (0.027) | 0.919 | 0.973 (0.050) | 0.744 |
| **Se** | 0.950 (0.112) | 0.846 | 0.900 (0.137) | 0.359 |
| **Sp** | 0.862 (0.126) | 0.868 | 0.969 (0.042) | 0.987 |
| **PD vs MSA-P** | **Training** | **Test** | **Training** | **Test** |
| **BA** | 0.800 (0.083) | 0.741 | 0.804 (0.116) | 0.710 |
| **AUC** | 0.946 (0.034) | 0.823 | 0.969 (0.069) | 0.839 |
| **Se** | 0.600 (0.167) | 0.816 | 0.708 (0.138) | 0.553 |
| **Sp** | 1.000 (0.000) | 0.667 | 0.900 (0.224) | 0.867 |
| **PD vs MSA-C** | **Training** | **Test** | **Training** | **Test** |
| **BA** | 0.915 (0.074) | 0.881 | 0.923 (0.038) | 0.802 |
| **AUC** | 1.000 (0.000) | 0.945 | 1.000 (0.000) | 0.932 |
| **Se** | 0.831 (0.148) | 0.829 | 0.846 (0.077) | 0.671 |
| **Sp** | 1.000 (0.000) | 0.933 | 1.000 (0.000) | 0.933 |
| **PD vs atypical parkinsonism** | **Training** | **Test** | **Training** | **Test** |
| **BA** | 0.826 (0.103) | 0.769 | 0.835 (0.088) | 0.775 |
| **AUC** | 0.945 (0.051) | 0.866 | 0.942 (0.033) | 0.861 |
| **Se** | 0.831 (0.158) | 0.908 | 0.892 (0.069) | 0.895 |
| **Sp** | 0.822 (0.169) | 0.631 | 0.778 (0.208) | 0.655 |
| **PSP vs MSA-C** | **Training** | **Test** | **Training** | **Test** |
| **BA** | 0.850 (0.105) | 0.928 | 0.858 (0.091) | 0.756 |
| **AUC** | 0.983 (0.037) | 0.978 | 0.967 (0.075) | 0.950 |
| **Se** | 0.900 (0.137) | 0.923 | 0.850 (0.224) | 0.513 |
| **Sp** | 0.800 (0.183) | 0.933 | 0.867 (0.183) | 1.000 |
| **PSP vs MSA-P** | **Training** | **Test** | **Training** | **Test** |
| **BA** | 0.660 (0.222) | 0.795 | 0.650 (0.224) | 0.56 |
| **AUC** | 0.640 (0.251) | 0.956 | 0.680 (0.259) | 0.834 |
| **Se** | 0.720 (0.228) | 0.923 | 0.600 (0.316) | 0.154 |
| **Sp** | 0.600 (0.418) | 0.667 | 0.700 (0.447) | 0.967 |
| **MSA-C vs MSA-P** | **Training** | **Test** | **Training** | **Test** |
| **BA** | 0.733 (0.181) | 0.617 | 0.567 (0.149) | 0.633 |
| **AUC** | 0.833 (0.204) | 0.833 | 0.667 (0.118) | 0.811 |
| **Se** | 0.800 (0.274) | 0.367 | 0.600 (0.224) | 0.400 |
| **Sp** | 0.667 (0.236) | 0.867 | 0.533 (0.298) | 0.867 |

HC: healthy controls; PD: Parkinson's disease; PSP: Progressive supranuclear palsy; MSA-C: cerebellar variant of multiple system atrophy; MSA-P: parkinsonian variant of multiple system atrophy; DTI: diffusion tensor imaging; BA: balanced accuracy; Se: sensitivity; Sp: specificity; SVM: support-vector machine

In the replication cohort, when using volumetric data solely, the BAs were slightly lower for the classifications of PD versus PSP, PD versus MSA-P, PD versus MSA-C, PD versus atypical parkinsonian syndromes, in comparison with the training data set, ranging between 0.741 and 0.881, with AUCs between 0.823 and 0.945. By contrast, better accuracies were obtained in the replication cohort for the classifications of PSP versus MSA-P and PSP versus MSA-C (BAs of 0.795 versus 0.660 and 0.928 versus 0.850, respectively). Adding the DTI metrics decreased categorization performances in most cases (Table 2).

### Effect of the normalization using HCs in the subset of the replication cohort

In the subset of the replication cohort, after normalization using HCs, adding DTI metrics to volumetric data increased BAs and/or AUCs of most classifications, except for the classifications of PSP versus MSA-P and MSA-C versus MSA-P (supplementary Table S4).

Both methods of normalization were compared in the subset. Combining volumetric and DTI metrics significantly increased accuracy for the normalization by HCs as compared with normalization by the training cohort for differentiation between PD and PSP (AUCs: 0.833 versus 0.935, p = 0.01). There was no significant difference between the classifications of PD versus MSA-C (AUCs: 0.992 versus 0.998, p>0.05) and PSP versus MSA-C (AUCs: 0.982 versus 0.997, p>0.05) (Table 3).

**Table 3:**
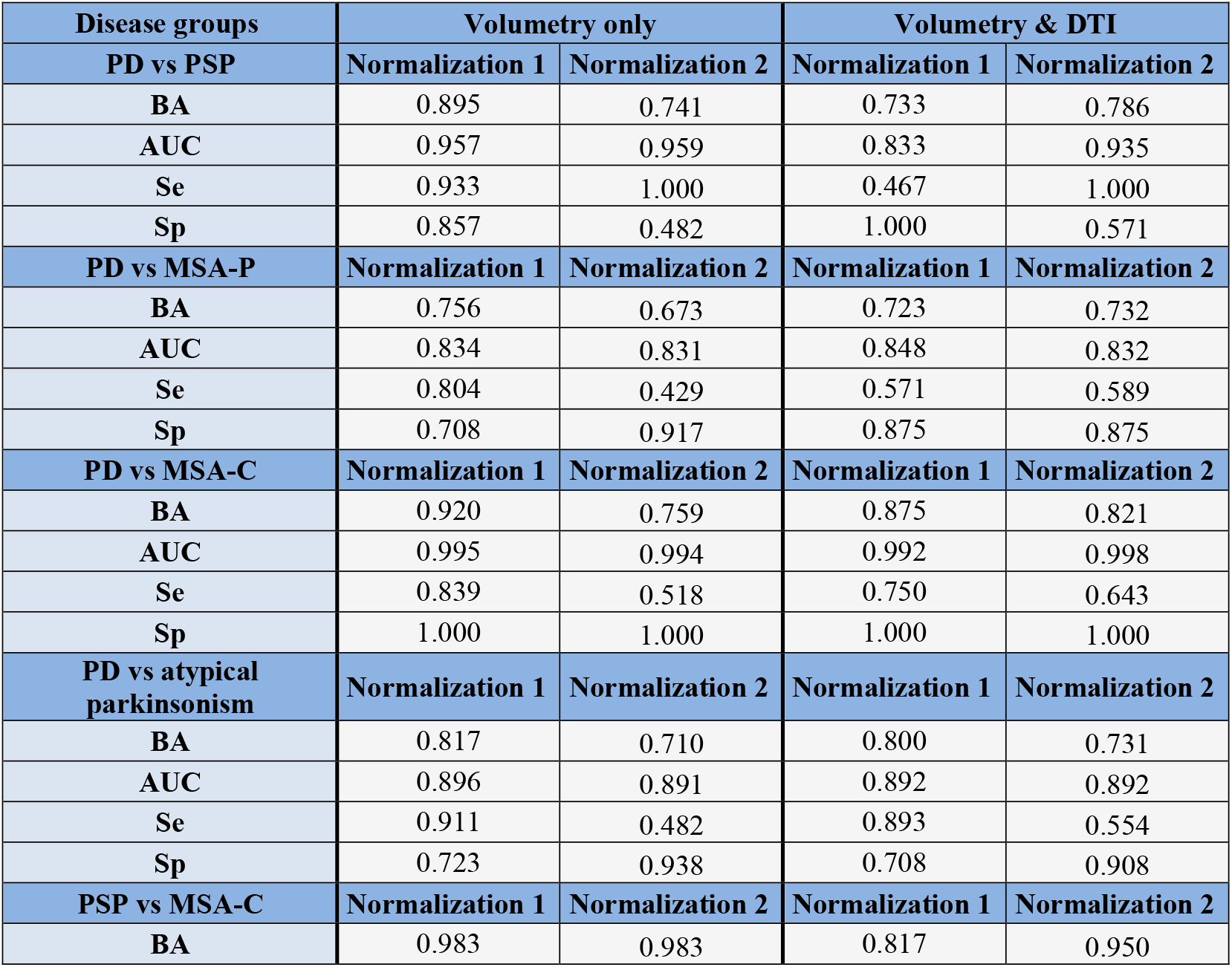
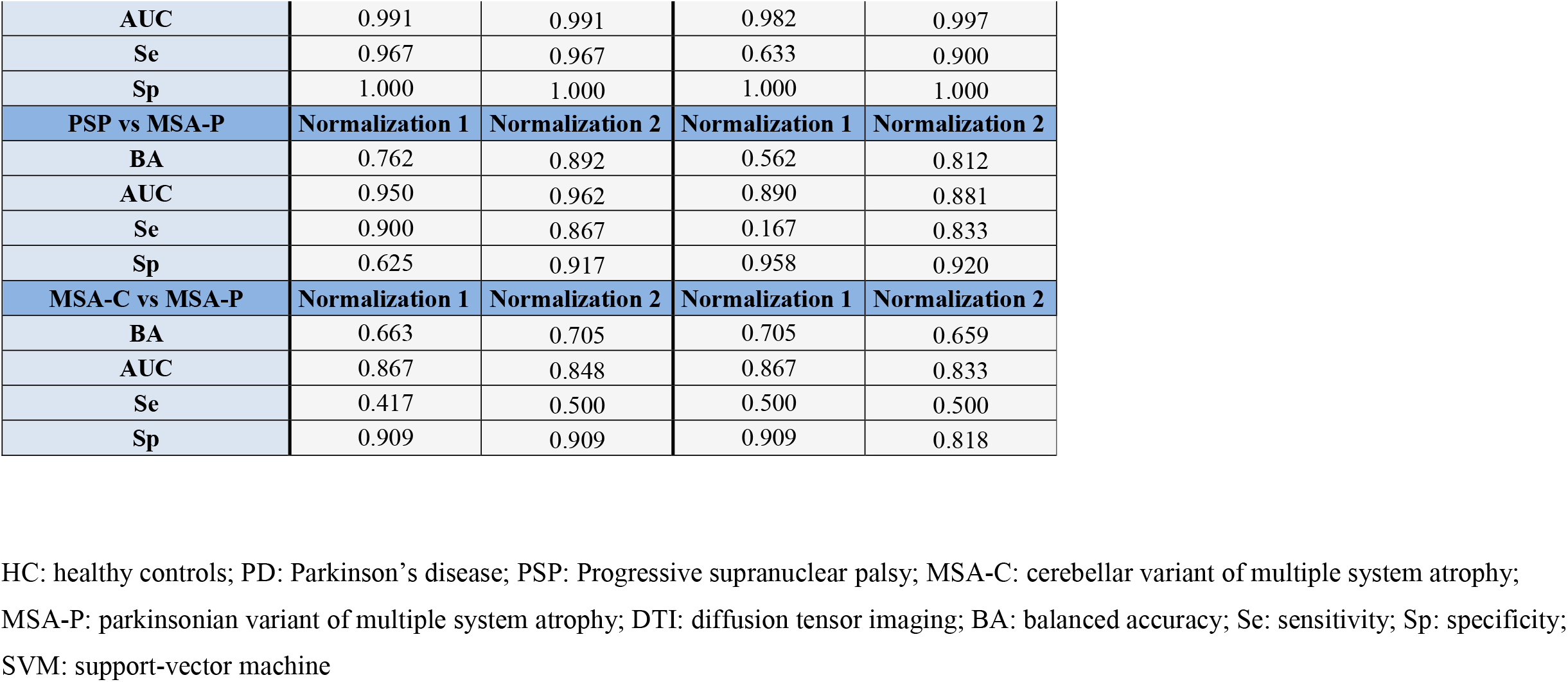
Performances of the linear SVM classification in the subset of the replication cohort after normalization using subjects of the training cohort (normalization 1) and normalization using healthy controls (normalization 2), for volumetric data and the combination of volumetric and DTI data.

| **Disease groups** | **Volumetry only** |  | **Volumetry & DTI** |  |
| --- | --- | --- | --- | --- |
| **PD vs PSP** | **Normalization 1** | **Normalization 2** | **Normalization 1** | **Normalization 2** |
| **BA** | 0.895 | 0.741 | 0.733 | 0.786 |
| **AUC** | 0.957 | 0.959 | 0.833 | 0.935 |
| **Se** | 0.933 | 1.000 | 0.467 | 1.000 |
| **Sp** | 0.857 | 0.482 | 1.000 | 0.571 |
| **PD vs MSA-P** | **Normalization 1** | **Normalization 2** | **Normalization 1** | **Normalization 2** |
| **BA** | 0.756 | 0.673 | 0.723 | 0.732 |
| **AUC** | 0.834 | 0.831 | 0.848 | 0.832 |
| **Se** | 0.804 | 0.429 | 0.571 | 0.589 |
| **Sp** | 0.708 | 0.917 | 0.875 | 0.875 |
| **PD vs MSA-C** | **Normalization 1** | **Normalization 2** | **Normalization 1** | **Normalization 2** |
| **BA** | 0.920 | 0.759 | 0.875 | 0.821 |
| **AUC** | 0.995 | 0.994 | 0.992 | 0.998 |
| **Se** | 0.839 | 0.518 | 0.750 | 0.643 |
| **Sp** | 1.000 | 1.000 | 1.000 | 1.000 |
| **PD vs atypical parkinsonism** | **Normalization 1** | **Normalization 2** | **Normalization 1** | **Normalization 2** |
| **BA** | 0.817 | 0.710 | 0.800 | 0.731 |
| **AUC** | 0.896 | 0.891 | 0.892 | 0.892 |
| **Se** | 0.911 | 0.482 | 0.893 | 0.554 |
| **Sp** | 0.723 | 0.938 | 0.708 | 0.908 |
| **PSP vs MSA-C** | **Normalization 1** | **Normalization 2** | **Normalization 1** | **Normalization 2** |
| **BA** | 0.983 | 0.983 | 0.817 | 0.950 |
| **AUC** | 0.991 | 0.991 | 0.982 | 0.997 |
| **Se** | 0.967 | 0.967 | 0.633 | 0.900 |
| **Sp** | 1.000 | 1.000 | 1.000 | 1.000 |
| **PSP vs MSA-P** | **Normalization 1** | **Normalization 2** | **Normalization 1** | **Normalization 2** |
| **BA** | 0.762 | 0.892 | 0.562 | 0.812 |
| **AUC** | 0.950 | 0.962 | 0.890 | 0.881 |
| **Se** | 0.900 | 0.867 | 0.167 | 0.833 |
| **Sp** | 0.625 | 0.917 | 0.958 | 0.920 |
| **MSA-C vs MSA-P** | **Normalization 1** | **Normalization 2** | **Normalization 1** | **Normalization 2** |
| **BA** | 0.663 | 0.705 | 0.705 | 0.659 |
| **AUC** | 0.867 | 0.848 | 0.867 | 0.833 |
| **Se** | 0.417 | 0.500 | 0.500 | 0.500 |
| **Sp** | 0.909 | 0.909 | 0.909 | 0.818 |

HC: healthy controls; PD: Parkinson's disease; PSP: Progressive supranuclear palsy; MSA-C: cerebellar variant of multiple system atrophy; MSA-P: parkinsonian variant of multiple system atrophy; DTI: diffusion tensor imaging; BA: balanced accuracy; Se: sensitivity; Sp: specificity; SVM: support-vector machine

### Weighting factors

The weighting factors are represented in Table 4 and Figure.

**Table 4:** Weighting factors extracted from the linear SVM training for the different group comparisons.

|  | PD vs PSP | PD vs MSA-P | PD vs MSA-C | PD vs atypical parkinsonism | PSP vs MSA-C | PSP vs MSA-P | MSA-C vs MSA-P |
| --- | --- | --- | --- | --- | --- | --- | --- |
| Midbrain | -1.00 | 0.18 | 0.55 | 1.00 | -0.02 | -0.60 | 0.39 |
| Pons | -0.34 | 0.30 | 1.00 | 0.80 | 0.73 | 0.25 | 1.00 |
| Midbrain to pons ratio | -0.04 | -0.78 | -1.00 | -0.60 | -1.00 | -1.00 | -0.93 |
| SCP | -0.38 | 0.15 | 0.55 | 0.64 | 0.34 | 0.10 | 0.53 |
| V3 | 0.99 | -0.25 | 0.32 | -0.63 | 1.00 | 1.00 | 0.70 |
| V4 | 1.00 | -0.88 | -0.31 | -1.00 | 0.14 | 0.12 | -0.24 |
| Cerebellum | -0.10 | 0.00 | 0.51 | 0.45 | 0.53 | 0.18 | 0.76 |
| Thalamus | -0.09 | -0.05 | 0.12 | 0.14 | -0.01 | -0.04 | -0.02 |
| Caudate | 0.63 | -0.04 | -0.01 | -0.32 | 0.28 | 0.51 | -0.12 |
| Putamen | -0.12 | 1.00 | 0.05 | 0.58 | 0.09 | 0.99 | -1.00 |
| Pallidum | -0.66 | 0.64 | 0.39 | 0.86 | 0.00 | 0.18 | -0.45 |
| Insula | 0.32 | -0.13 | 0.13 | -0.23 | 0.38 | 0.27 | 0.53 |
| Precentral | 0.50 | -0.11 | -0.16 | -0.30 | 0.14 | 0.40 | -0.13 |
| Midbrain FA | -0.43 | 0.05 | 0.53 | 0.50 | 0.22 | -0.11 | 0.39 |
| Pons FA | -0.28 | 0.28 | 0.54 | 0.43 | 0.33 | 0.22 | 0.25 |
| SCP FA | -0.87 | 0.34 | 0.59 | 0.90 | -0.09 | -0.51 | 0.22 |
| Putamen FA | 0.79 | -0.63 | -0.03 | -0.66 | 0.42 | 0.01 | 0.63 |
| Posteriorputamen FA | 0.63 | -0.53 | -0.02 | -0.49 | 0.33 | 0.13 | 0.28 |
| Pallidum FA | 0.19 | -0.29 | 0.31 | 0.01 | 0.41 | 0.13 | 0.39 |
| Thalamus FA | 0.23 | -0.16 | 0.20 | -0.12 | 0.18 | -0.31 | 0.55 |
| Caudate FA | 0.60 | -0.33 | 0.18 | -0.30 | 0.45 | 0.22 | 0.52 |
| Cerebellum FA | -0.11 | -0.08 | 0.57 | 0.24 | 0.38 | -0.14 | 0.80 |
| Insula FA | 0.68 | -0.35 | 0.26 | -0.31 | 0.52 | 0.24 | 0.67 |
| Precentral FA | 0.19 | -0.24 | 0.33 | 0.05 | 0.35 | -0.04 | 0.62 |
| Midbrain MD | 0.65 | -0.24 | 0.08 | -0.39 | 0.45 | 0.51 | 0.32 |
| Pons MD | 0.47 | -0.49 | -0.01 | -0.33 | 0.15 | -0.10 | 0.39 |
| SCP MD | 0.39 | -1.00 | 0.06 | -0.46 | 0.15 | -0.62 | 0.72 |
| **Putamen MD** | 0.33 | -0.12 | 0.19 | -0.15 | 0.35 | 0.36 | 0.35 |
| **Posteriorputamen MD** | 0.13 | -0.27 | 0.01 | -0.15 | 0.18 | 0.18 | 0.26 |
| **Pallidum MD** | 0.13 | 0.25 | 0.41 | 0.19 | 0.43 | 0.55 | 0.40 |
| **Thalamus MD** | 0.13 | 0.15 | 0.50 | 0.23 | 0.52 | 0.55 | 0.32 |
| **Caudate MD** | 0.44 | -0.23 | 0.35 | -0.20 | 0.47 | 0.33 | 0.36 |
| **Cerebellum MD** | 0.11 | -0.35 | 0.10 | 0.04 | 0.07 | -0.26 | 0.27 |
| **Insula MD** | 0.48 | -0.07 | 0.36 | -0.15 | 0.53 | 0.52 | 0.28 |
| **Precentral MD** | 0.10 | 0.26 | 0.56 | 0.20 | 0.48 | 0.47 | 0.31 |
| **Midbrain AD** | 0.36 | 0.06 | 0.18 | -0.09 | 0.41 | 0.63 | 0.12 |
| **Pons AD** | 0.27 | -0.35 | 0.01 | -0.17 | 0.07 | -0.11 | 0.23 |
| **SCP AD** | -0.20 | -0.67 | 0.26 | 0.07 | -0.02 | -0.85 | 0.59 |
| **Putamen AD** | 0.47 | -0.22 | 0.13 | -0.30 | 0.35 | 0.30 | 0.41 |
| **Pallidum AD** | 0.07 | 0.34 | 0.51 | 0.31 | 0.47 | 0.60 | 0.41 |
| **Thalamus AD** | 0.07 | 0.21 | 0.52 | 0.30 | 0.51 | 0.53 | 0.31 |
| **Caudate AD** | 0.45 | -0.23 | 0.38 | -0.18 | 0.49 | 0.33 | 0.37 |
| **Cerebellum AD** | 0.02 | -0.31 | 0.20 | 0.11 | 0.09 | -0.31 | 0.39 |
| **Insula AD** | 0.55 | -0.06 | 0.40 | -0.15 | 0.57 | 0.55 | 0.33 |
| **Precentral AD** | 0.01 | 0.33 | 0.67 | 0.35 | 0.51 | 0.43 | 0.39 |
| **Midbrain RD** | 0.67 | -0.29 | -0.11 | -0.51 | 0.40 | 0.64 | 0.39 |
| **Pons RD** | 0.45 | -0.27 | -0.19 | -0.31 | 0.14 | 0.22 | 0.27 |
| **SCP RD** | 0.59 | -0.87 | -0.09 | -0.61 | 0.39 | 0.22 | 0.74 |
| **Putamen RD** | 0.14 | 0.05 | 0.24 | 0.05 | 0.44 | 0.31 | 0.64 |
| **Pallidum RD** | -0.04 | 0.41 | 0.59 | 0.44 | 0.56 | 0.54 | 0.53 |
| **Thalamus RD** | 0.12 | 0.29 | 0.41 | 0.22 | 0.55 | 0.62 | 0.41 |
| **Caudate RD** | 0.46 | -0.10 | 0.29 | -0.23 | 0.52 | 0.48 | 0.34 |
| **Cerebellum RD** | 0.34 | -0.38 | -0.12 | -0.27 | 0.01 | -0.16 | 0.07 |
| **Insula RD** | 0.53 | -0.14 | 0.30 | -0.30 | 0.59 | 0.54 | 0.73 |
| **Precentral RD** | 0.27 | 0.14 | 0.56 | 0.00 | 0.62 | 0.66 | 0.60 |

Weighting factors were scaled to a range of -1 to +1. The higher the absolute value, the bigger the contribution of the feature. When the coefficient was positive, the algorithm favored the first disease group if the value of the feature was high or the second group if the value was low. Conversely, when the coefficient was negative, the algorithm favored the second group if the value of the feature was high, or the first group if the value was low. Positive values are highlighted in shades of red while negative values are highlighted in shades of blue.
HC: healthy controls; PD: Parkinson's disease; PSP: Progressive supranuclear palsy; MSA-C: cerebellar variant of multiple system atrophy; MSA-P: parkinsonian variant of multiple system atrophy; AD: axial diffusivity; FA: fractional anisotropy; MD: mean diffusivity; RD: radial diffusivity; SCP: superior cerebellar peduncles; V3: third ventricle; V4: fourth ventricle.

The best features for the differentiation between PD and PSP were midbrain (1), third ventricle (−1) and fourth ventricle (−1) volumes, followed by FA in the SCP (−0.87) and the putamen (0.79).

For discriminating subjects with MSA-C, the most relevant features were midbrain to pons ratio (−0.93 to −1), pons volume (−0.78 to 1), cerebellum volume (0.51 to 0.76). Moreover MSA-C was well differentiated from MSA-P using putamen volume (−1), V3 volume (0.70), FA in the cerebellum (0.80), MD and RD in the SCP (0.72 and 0.74, respectively), and FA and RD in the insula (0.67 and 0.73, respectively). For the classification of PD versus MSA-C, FA in the midbrain (0.53), the pons (0.54) and the SCP (0.59) were also relevant. DTI metrics in the brainstem and the cerebellum did not efficiently differentiate between PSP and MSA-C.

MSA-P differentiation relied mostly on putamen volume (versus PSP: 0.99, versus PD: 1, versus MSA-C: −1) and midbrain to pons ratio (−0.93 to −1). FA, MD and AD in the SCP (−0.51, −0.62, −0.85) were relevant for the differentiation of PSP and MSA-P, unlike diffusion in the putamen.

## Discussion

Our study demonstrated the feasibility of an automated classification of parkinsonian syndromes in a clinical setting using a machine learning algorithm developed in a research environment. To date, we provide the third largest cohort ever analyzed.^19,25^. The originality of the study is that the algorithm was tested on a large independent replication cohort composed of patients recruited in a movement disorders clinic and scanned across four different scanners in a Neuroradiology department, in the frame of their routine diagnostic workup. Patients in the replication cohort had a shorter disease duration than those in the training cohort, making the algorithm more robust for the differentiation of patients with early to moderately advanced parkinsonism. A further strength is that patients with both parkinsonian and cerebellar subtypes of MSA were included in addition to PD and PSP, which was done in only two previous studies.^19,23^

Our results were consistent with previous publications showing high accuracy in the machine-learning-based differentiation of parkinsonian syndromes, as BAs ranged between 69 and 89% in the study by Huppertz et al.,^19^ and AUCs were respectively greater than 93% and 95% in the studies by Archer et al.,^25^ and Scherfler et al.^18^ Nevertheless, the performances are difficult to compare across studies given differences regarding study designs in terms of input data and group diseases, classification problems (binary or multiclass approaches) and performance indices used. It has been shown that morphometric measurements are robust MRI markers for the discrimination of parkinsonism.^18–20^ This finding was confirmed by our study in both the training and replication cohorts, though the accuracy was lower for the classification of PSP versus MSA-P and MSA-P versus MSA-C. These results may be explained in our study by the relative small number of MSA-P and MSA-C patients in the training cohort that might have reduced the performances of the algorithm. Moreover, MSA-P and MSA-C patients may often have overlapping features, a clinical and brain imaging continuum existing between both variants.^5^ In our study, patients exhibiting both patterns of MSA were pooled with MSA-P patients as isolating a group with putaminal abnormalities appeared to be more relevant.

In the training cohort, including DTI metrics improved BAs for most group comparisons (except for the classifications of PSP versus MSA-P and MSA-C versus MSA-P), the best differentiation being achieved for the classifications of PD versus PSP and PD versus MSA-C. These results suggest that DTI metrics standardized in a research protocol provide additional information useful for the group differentiation. Conversely, in the replication cohort, adding DTI metrics decreased categorization performances in most cases. The heterogeneity of data in the replication cohort, which comprised images acquired using distinct MRI systems and variable acquisition parameters, probably accounted for such a decrease in categorization accuracy. More specifically, DTI image resolution was isotropic with no inter-slice gap in the training cohort, whereas it was anisotropic with a variable inter-slice gap in the replication cohort. In contrast, the structural T1-weighted images had a very similar isotropic resolution (close to1 mm^3^) in all cohorts. In the replication cohort, the inconsistent DTI image resolution across subjects scanned using distinct MRI systems could have resulted in the inconsistent sampling of tissue diffusion properties, thus contributing to decreasing the performance of machine-learning-based patient categorization when combining DTI data with isotropic structural data. It is probable that such an effect was not observed in the training cohort because both structural and diffusion tissue properties were sampled isotropically.

To evaluate the scanner effect, a second analysis was performed on a subset of the replication cohort using a normalization by HCs values. BAs and/or AUCs increased when combining volume and diffusion except for the classifications of PSP versus MSA-P and MSA-C versus MSA-P. When comparing both methods of normalization in the subset, the performances only slightly increased for the classification between PD and PSP, showing the difficulty to manage the scanner effect. Improvement in DTI acquisition parameters, especially standardization of the resolution and slice gap, may improve classification performances as suggested previously. ^25^

In line with previous pathological and imaging studies, the best features for the differentiation between PD and PSP were midbrain and third ventricle volumes ^6,7,13,17,28,29^ and FA in the SCP.^17,29–31^ The putamen volume was highly discriminant between MSA-P and PD, PSP and MSA-C patients in our study, MSA-P being characterized by a prominent putamen atrophy.^14,17,28,30^ DTI metrics in the SCP were also relevant for the differentiation of MSA-P and PSP as shown in previous studies.^14,17,30–32^

Unlike atrophy, diffusion in the whole putamen and posterior putamen did not significantly contribute to the differentiation of MSA-P versus other disease groups in our study. Diffusivity in the posterior putamen was shown to be highly affected in MSA-P versus HCs and PD patients,^33–35^ with higher iron depositions.^36,37^ Conversely, several studies have demonstrated overlapping values in the entire putamen between PSP and MSA-P patients regarding diffusivity ^32,38,39^ and relaxometry.^39,40^ In one study, ADC values were increased in the posterior putamen in MSA-P patients compared to PSP patients.^35^ In our study, echo planar imaging distortions seen on DTI images and due to susceptibility artifacts may have resulted in the imperfect overlay between putamen masks derived from T1-weighted images and diffusion-weighted images. Therefore, co-registration inaccuracies have likely reduced the accuracy of the diffusion measurements and the performance of the algorithm.

One of the limitations of our study was the lack of neuropathological confirmation, which is intrinsic to most neurodegenerative studies. For now, the clinical follow-up is the only diagnostic “gold standard” we can rely on. In our institution, most patients are followed up by experts in movement disorders with a systematic visit two years after the first consultation, allowing to improve clinical diagnostic accuracy. Second, there was a large heterogeneity between DTI protocols, which indeed corresponds to clinical environment. In this context, it appears important to standardize DTI protocols across scanners and centers in order to improve categorization performances. Future work could involve calculating free-water and free-water-corrected measurements in clinical protocols for improved classification accuracy.^25^

In conclusion, our study showed that an automated categorization of parkinsonian syndromes was applicable to patients with early to moderately advanced parkinsonism recruited in a clinical environment, despite the variability in scanners and acquisition parameters, volumetry being a robust discriminative biomarker. Medical centers could benefit from such an approach in order to increase diagnostic accuracy and patient management. Implementing a machine learning algorithm in the clinical workflow of a Neuroradiology department may thus be relevant to help clinicians improve diagnosis of parkinsonism.

## Data Availability

I declare that all data referred to in the manuscript and note links below are available

## Authors’ Roles

Authors' Roles
| Name | Location | Role | Contribution |
| --- | --- | --- | --- |
| Lydia Chougar | Sorbonne Université,<br>ICM, Paris, France | Author | Designed and conceptualized the study; analyzed of the data; drafted the manuscript for intellectual content |
| Johann Faouzi | ARAMIS, ICM,<br>Paris, France | Author | Designed and conceptualized the study; analyzed of the data; revised the manuscript for intellectual content |
| Nadya Pyatigorskaya | Sorbonne Université,<br>ICM, Paris, France | Author | Designed and conceptualized study; revised the manuscript for intellectual content |
| Rahul Gaurav | CENIR, ICM, Paris,<br>France | Author | Data analysis, revision of manuscript |
| Emma Biondetti | CENIR, ICM, Paris,<br>France | Author | Data analysis, revision of manuscript |
| Marie Villotte | Université Denis<br>Diderot, Paris,<br>France | Author | Data collection and analysis, revision of manuscript |
| Romain Valabregue | CENIR, ICM, Paris,<br>France | Author | Data analysis, revision of manuscript |
| Jean-Christophe Corvol | CIC, ICM, Paris,<br>France | Author | Revision of manuscript |
| Alexis Brice | ICM, Paris, France | Author | Revision of manuscript |
| Louise-Laure Mariani | CIC, ICM, Paris,<br>France | Author | Revision of manuscript |
| Florence Cormier | Pitié Salpêtrière,<br>APHP, Paris, France | Author | Revision of manuscript |
| Marie Vidailhet | Sorbonne Université,<br>ICM, Paris, France | Author | Designed and conceptualized the study; revised the manuscript for intellectual content |
| Gwendoline Dupont | Université de<br>Bourgogne, Dijon,<br>France | Author | Data collection, revision of manuscript |
| Ines Piot | Pitié Salpêtrière,<br>APHP, Paris, France | Author | Data collection, revision of manuscript |
| David Grabli | Pitié Salpêtrière,<br>APHP, Paris, France | Author | Designed and conceptualized study; revised the manuscript for intellectual content |
| Christine Payan | Pitié Salpêtrière,<br>APHP, Paris, France | Author | Data collection, revision of manuscript |
| Olivier Colliot | ARAMIS, ICM,<br>Paris, France | Author | Designed and conceptualized the study; analyzed of the data; revised the manuscript for intellectual content |
| Bertrand Degos | APHP, Avicennes,<br>Paris, France | Author | Designed and conceptualized<br>study; revised the manuscript for<br>intellectual content |
| Stéphane Lehericy | Sorbonne Université,<br>ICM, Paris, France | Author | Designed and conceptualized<br>study; revised the manuscript for<br>intellectual content |

## Figure legend

### Contribution of each region of interest for the group classification using volumetry

A color shade was attributed to the weighting factor corresponding to each brain region depending on its relevance for the group differentiation. Weighting factors were rescaled to the range of 0 to 1.

The most relevant regions were the midbrain, the third ventricle and the fourth ventricle for the classification of PD versus PSP, the putamen and the fourth ventricle for PD versus MSA-P, the pons for PD versus MSA-C, the third ventricle and the pons for PSP versus MSA-C, the putamen and the third ventricle for PSP versus MSA-P, the putamen and the pons for MSA-P versus MSA-C.

HC: healthy controls; PD: Parkinson’s disease; PSP: Progressive supranuclear palsy; MSA-C: cerebellar variant of multiple system atrophy; MSA-P: parkinsonian variant of multiple system atrophy

## Supplementary tables

**Table S1:** Magnetic resonance imaging acquisition parameters.

| Machine | 3T TRIO |  | 1.5T OPTIMA | 3T SIGNA | 3T SKYRA | 3T PET/MR |
| --- | --- | --- | --- | --- | --- | --- |
| n | 179 |  | 32 | 79 | 84 | 7 |
| Magnetic field | 3T |  | 1.5T | 3T | 3T | 3T |
| Vendor | Siemens |  | GE | GE | Siemens | GE |
| Head coil | 32 |  | 32 | 32 | 64 | 32 |
| T1-w sequences |  |  |  |  |  |  |
| TE (ms) | 2,94 |  | 1 | 2,788 | 2,34 | 3,06 |
| TR (ms) | 2200 |  | 9,356 | 6,564 | 2100 | 7,208 |
| TI (ms) | 900 |  | 300 | 400 | 900 | 400 |
| Flip angle | 10 |  | 15° | 11° | 8° | 11° |
| Voxel size (mm) | 1x1x1 |  | 0.9x0.9x1 | 1x1x1 | 0.9x0.9x0.9 | 1x1x1 |
| DTI | Protocol 1 | Protocol 2 |  |  |  |  |
| Number, range of b-values | 2, 0-1000 | 2, 0-1500 | 2, 0-1000 | 2, 0-1000 | 2, 0-1000 | 2, 0-1000 |
| Directions | 64 | 60 | 15 | 15 | 30 | 30 |
| TE (ms) | 101 | 89 | 91,8 | 91,2 | 77 | 95 |
| TR (ms) | 14000 | 12000 | 9000 | 17000 | 3800 | 11500 |
| Flip angle | 90° | 90° | 90° | 90° | 90° | 90° |
| Voxel size (mm) | 2x2x2 | 1.7x1.7x1.7 | 1.0938x1.0938x3 | 1x1x2.5 | 2x2x2.6 | 1.25x1.25x2.5 |
| Slice gap | 0 | 0 | 0 | 0 | 0,6 | 0 |

DTI: Diffusion tensor imaging; n: number of patients scanned with a given MRI system; TE: echo time; TR: repetition time; T1-w: T1-weighted; GE: General Electrics Medical Systems
Two DTI protocols were acquired using the 3T TRIO MRI system.

**Table S2:**
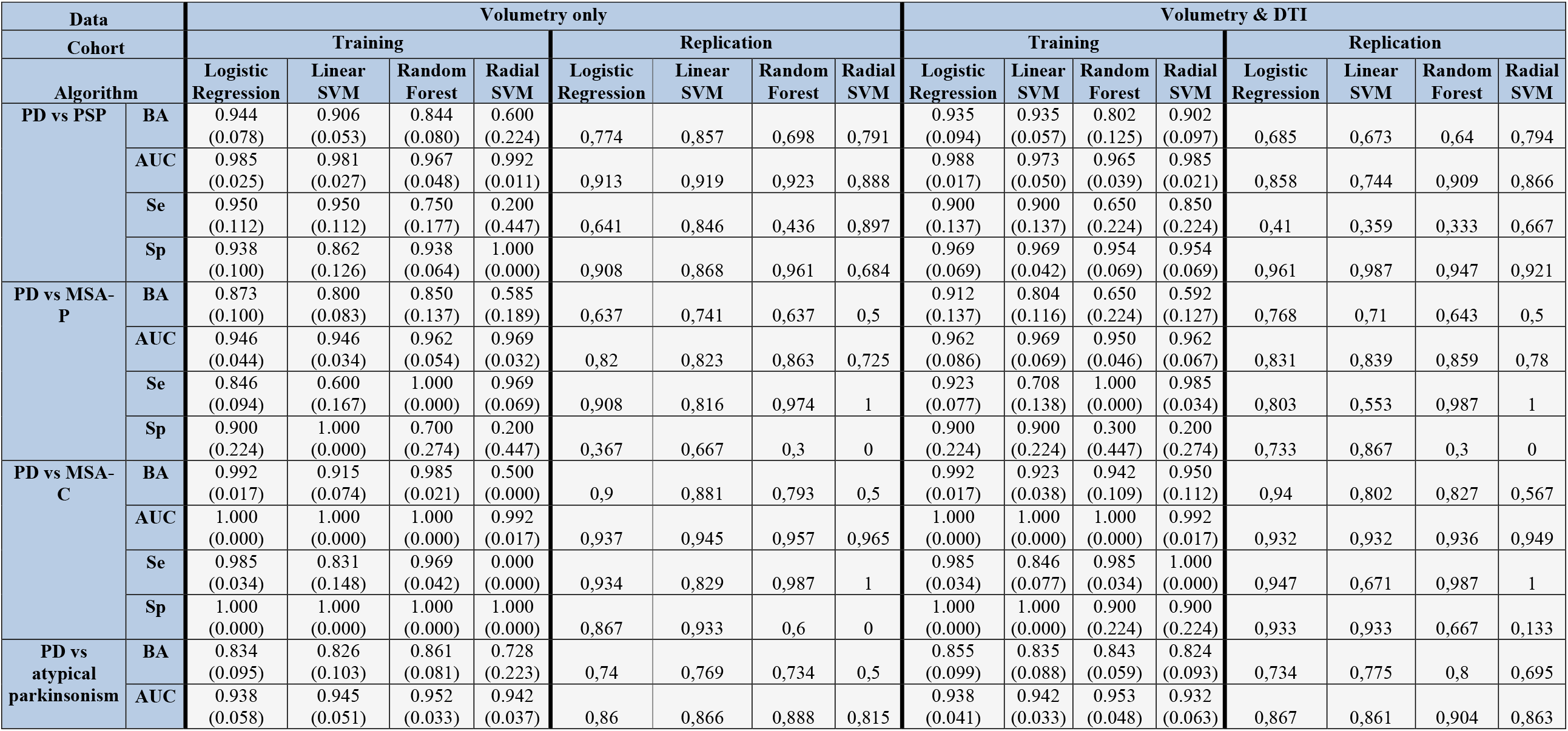
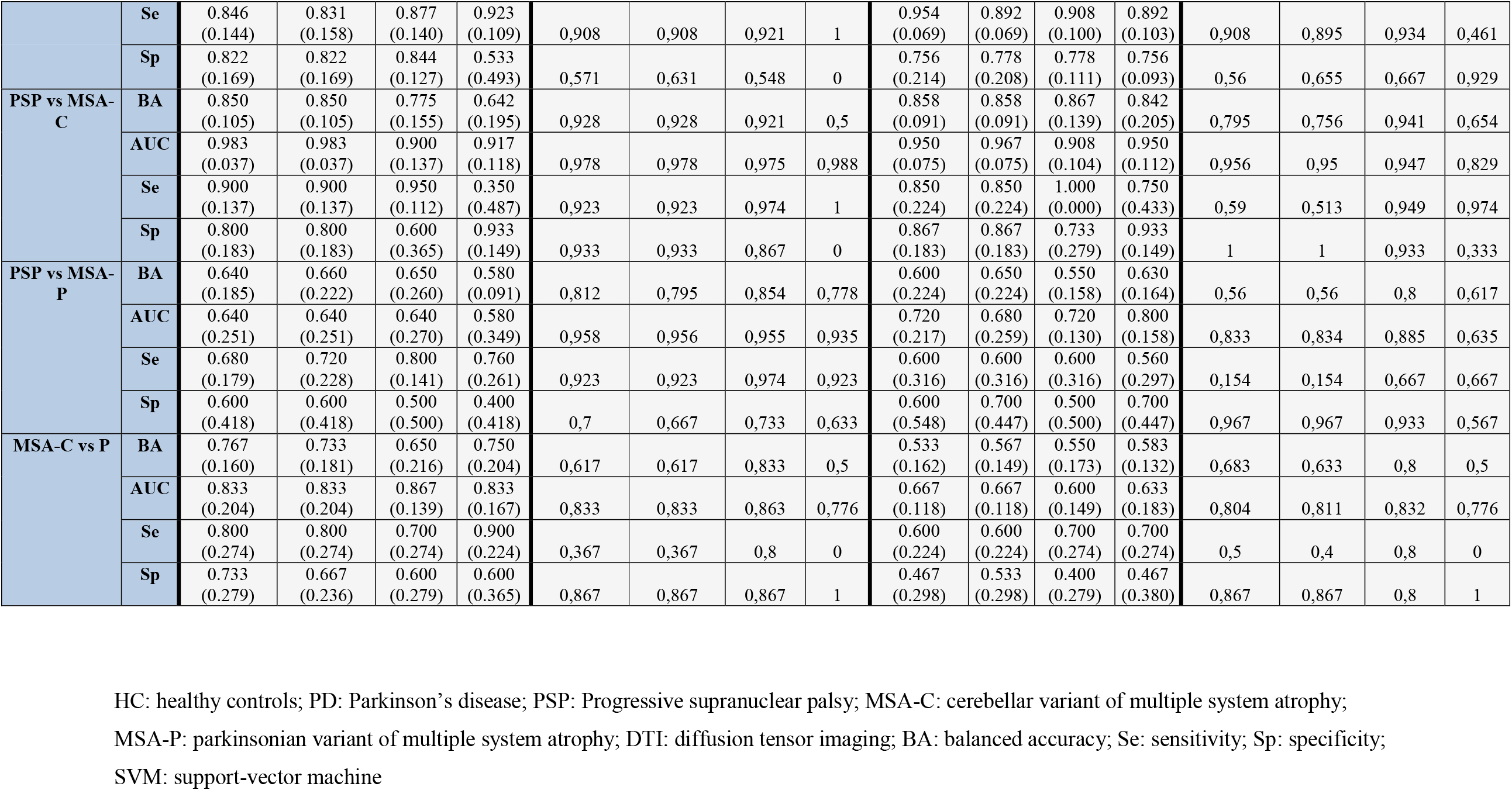
Performances of the four machine learning algorithms in the training cohort and in the whole replication cohort after normalization using subjects values in the training cohort for volumetric data and for the combination of volumetric and DTI metrics.

**Table S3:** Performances of the four machine learning algorithms in the training cohort and in the whole replication cohort after normalization using HCs values in each cohort for volumetric data and for the combination of volumetric and DTI metrics.

| Data |  | Volumetry only |  |  |  |  |  |  |  | Volumetry & DTI |  |  |  |  |  |  |  |
| --- | --- | --- | --- | --- | --- | --- | --- | --- | --- | --- | --- | --- | --- | --- | --- | --- | --- |
| Cohort |  | Training |  |  |  | Replication |  |  |  | Training |  |  |  | Replication |  |  |  |
| Algorithm |  | Logistic Regression | Linear SVM | Random Forest | Radial SVM | Logistic Regression | Linear SVM | Random Forest | Radial SVM | Logistic Regression | Linear SVM | Random Forest | Radial SVM | Logistic Regression | Linear SVM | Random Forest | Radial SVM |
| PD vs PSP | BA | 0.952<br>(0.073) | 0.790<br>(0.157) | 0.902<br>(0.063) | 0.700<br>(0.274) | 0.904 | 0.741 | 0.79 | 0.876 | 0.927<br>(0.055) | 0.777<br>(0.050) | 0.892<br>(0.096) | 0.735<br>(0.215) | 0.871 | 0.786 | 0.781 | 0.865 |
|  | AUC | 0.981<br>(0.027) | 0.954<br>(0.075) | 0.979<br>(0.029) | 0.992<br>(0.011) | 0.958 | 0.959 | 0.939 | 0.95 | 0.981<br>(0.027) | 0.977<br>(0.021) | 0.988<br>(0.026) | 0.985<br>(0.021) | 0.945 | 0.935 | 0.951 | 0.936 |
|  | Se | 0.950<br>(0.112) | 0.950<br>(0.112) | 0.850<br>(0.137) | 0.400<br>(0.548) | 0.933 | 1 | 0.633 | 0.967 | 0.900<br>(0.137) | 1.000<br>(0.000) | 0.800<br>(0.209) | 0.500<br>(0.468) | 0.867 | 1 | 0.633 | 0.767 |
|  | Sp | 0.954<br>(0.069) | 0.631<br>(0.213) | 0.954<br>(0.069) | 1.000<br>(0.000) | 0.875 | 0.482 | 0.946 | 0.786 | 0.954<br>(0.069) | 0.554<br>(0.100) | 0.985<br>(0.034) | 0.969<br>(0.069) | 0.875 | 0.571 | 0.929 | 0.964 |
| PD vs MSA-P | BA | 0.881<br>(0.117) | 0.708<br>(0.058) | 0.800<br>(0.112) | 0.585<br>(0.189) | 0.804 | 0.673 | 0.595 | 0.5 | 0.912<br>(0.112) | 0.719<br>(0.121) | 0.650<br>(0.224) | 0.777<br>(0.194) | 0.747 | 0.732 | 0.574 | 0.625 |
|  | AUC | 0.946<br>(0.034) | 0.923<br>(0.067) | 0.969<br>(0.032) | 0.954<br>(0.063) | 0.85 | 0.831 | 0.868 | 0.827 | 0.962<br>(0.086) | 0.931<br>(0.092) | 0.942<br>(0.064) | 0.931<br>(0.134) | 0.851 | 0.832 | 0.813 | 0.804 |
|  | Se | 0.862<br>(0.034) | 0.415<br>(0.117) | 1.000<br>(0.000) | 0.969<br>(0.069) | 0.857 | 0.429 | 0.982 | 1 | 0.923<br>(0.000) | 0.538<br>(0.094) | 1.000<br>(0.000) | 0.954<br>(0.042) | 0.911 | 0.589 | 0.982 | 1 |
|  | Sp | 0.900<br>(0.224) | 1.000<br>(0.000) | 0.600<br>(0.224) | 0.200<br>(0.447) | 0.75 | 0.917 | 0.208 | 0 | 0.900<br>(0.224) | 0.900<br>(0.224) | 0.300<br>(0.447) | 0.600<br>(0.418) | 0.583 | 0.875 | 0.167 | 0.25 |
| PD vs MSA-C | BA | 1.000<br>(0.000) | 0.808<br>(0.061) | 0.969<br>(0.042) | 0.677<br>(0.243) | 0.982 | 0.759 | 0.955 | 0.5 | 1.000<br>(0.000) | 0.908<br>(0.070) | 0.892<br>(0.131) | 0.750<br>(0.250) | 0.991 | 0.821 | 0.991 | 0.909 |
|  | AUC | 1.000<br>(0.000) | 0.992<br>(0.017) | 1.000<br>(0.000) | 0.977<br>(0.052) | 0.995 | 0.994 | 0.99 | 0.987 | 1.000<br>(0.000) | 0.992<br>(0.017) | 0.992<br>(0.017) | 1.000<br>(0.000) | 0.995 | 0.998 | 0.995 | 0.994 |
|  | Se | 1.000<br>(0.000) | 0.615<br>(0.122) | 0.938<br>(0.084) | 0.354<br>(0.485) | 0.964 | 0.518 | 0.911 | 1 | 1.000<br>(0.000) | 0.815<br>(0.140) | 0.985<br>(0.034) | 0.600<br>(0.548) | 0.982 | 0.643 | 0.982 | 1 |
|  | Sp | 1.000<br>(0.000) | 1.000<br>(0.000) | 1.000<br>(0.000) | 1.000<br>(0.000) | 1 | 1 | 1 | 0 | 1.000<br>(0.000) | 1.000<br>(0.000) | 0.800<br>(0.274) | 0.900<br>(0.224) | 1 | 1 | 1 | 0,818 |
| PD vs atypical parkinsonism | BA | 0.827<br>(0.100) | 0.821<br>(0.095) | 0.831<br>(0.096) | 0.803<br>(0.184) | 0,843 | 0,71 | 0,816 | 0,827 | 0.843<br>(0.102) | 0.827<br>(0.028) | 0.843<br>(0.103) | 0.747<br>(0.232) | 0,829 | 0,731 | 0,853 | 0,833 |
|  | AUC | 0.938<br>(0.049) | 0.935<br>(0.053) | 0.955<br>(0.044) | 0.952<br>(0.046) | 0,897 | 0,891 | 0,904 | 0,906 | 0.940<br>(0.034) | 0.916<br>(0.048) | 0.950<br>(0.040) | 0.937<br>(0.061) | 0,902 | 0,892 | 0,898 | 0,902 |
|  | Se | 0.877<br>(0.117) | 0.708<br>(0.274) | 0.862<br>(0.138) | 0.938<br>(0.084) | 0,839 | 0,482 | 0,893 | 0,839 | 0.908<br>(0.084) | 0.677<br>(0.100) | 0.908<br>(0.100) | 0.938<br>(0.084) | 0,857 | 0,554 | 0,875 | 0,804 |
|  | Sp | 0.778<br>(0.176) | 0.933<br>(0.099) | 0.800<br>(0.165) | 0.667<br>(0.393) | 0,846 | 0,938 | 0,738 | 0,815 | 0.778<br>(0.208) | 0.978<br>(0.050) | 0.778<br>(0.222) | 0.556<br>(0.509) | 0,8 | 0,908 | 0,831 | 0,862 |
| PSP vs MSA-C | BA | 0.867<br>(0.139) | 0.850<br>(0.105) | 0.842<br>(0.119) | 0.500<br>(0.000) | 0,983 | 0,983 | 0,967 | 0,5 | 0.800<br>(0.183) | 1.000<br>(0.000) | 0.800<br>(0.183) | 0.500<br>(0.000) | 0,967 | 0,95 | 0,938 | 0,5 |
|  | AUC | 0.983<br>(0.037) | 0.983<br>(0.037) | 0.933<br>(0.109) | 0.967<br>(0.046) | 0,991 | 0,991 | 0,991 | 0,991 | 1.000<br>(0.000) | 1.000<br>(0.000) | 0.892<br>(0.130) | 1.000<br>(0.000) | 0,997 | 0,997 | 0,97 | 0,991 |
|  | Se | 1.000<br>(0.000) | 0.900<br>(0.137) | 0.950<br>(0.112) | 0.000<br>(0.000) | 0,967 | 0,967 | 0,933 | 1 | 1.000<br>(0.000) | 1.000<br>(0.000) | 1.000<br>(0.000) | 0.000<br>(0.000) | 0,933 | 0,9 | 0,967 | 1 |
|  | Sp | 0.733<br>(0.279) | 0.800<br>(0.183) | 0.733<br>(0.279) | 1.000<br>(0.000) | 1 | 1 | 1 | 0 | 0.600<br>(0.365) | 1.000<br>(0.000) | 0.600<br>(0.365) | 1.000<br>(0.000) | 1 | 1 | 0,909 | 0 |
| PSP vs MSA-P | BA | 0.660<br>(0.248) | 0.730<br>(0.175) | 0.630<br>(0.286) | 0.560<br>(0.134) | 0,875 | 0,892 | 0,863 | 0,5 | 0.700<br>(0.170) | 0.700<br>(0.154) | 0.660<br>(0.233) | 0.610<br>(0.175) | 0,775 | 0,812 | 0,733 | 0,771 |
|  | AUC | 0.720<br>(0.192) | 0.780<br>(0.228) | 0.600<br>(0.292) | 0.600<br>(0.255) | 0,946 | 0,962 | 0,968 | 0,967 | 0.920<br>(0.084) | 0.920<br>(0.084) | 0.620<br>(0.286) | 0.900<br>(0.071) | 0,885 | 0,881 | 0,856 | 0,881 |
|  | Se | 0.720<br>(0.228) | 0.760<br>(0.167) | 0.760<br>(0.167) | 0.920<br>(0.179) | 0,833 | 0,867 | 0,933 | 0 | 0.800<br>(0.200) | 0.800<br>(0.141) | 0.920<br>(0.110) | 0.920<br>(0.110) | 0,8 | 0,833 | 0,967 | 0,833 |
|  | Sp | 0.600<br>(0.418) | 0.700<br>(0.274) | 0.500<br>(0.500) | 0.200<br>(0.447) | 0,917 | 0,917 | 0,792 | 1 | 0.600<br>(0.418) | 0.600<br>(0.418) | 0.400<br>(0.418) | 0.300<br>(0.447) | 0,75 | 0,792 | 0,5 | 0,708 |
| MSA-C vs P | BA | 0.767<br>(0.181) | 0.900<br>(0.137) | 0.650<br>(0.216) | 0.800<br>(0.192) | 0,784 | 0,705 | 0,705 | 0,5 | 0.700<br>(0.173) | 0.733<br>(0.181) | 0.517<br>(0.109) | 0.833<br>(0.177) | 0,809 | 0,659 | 0,792 | 0,5 |
|  | AUC | 0.867<br>(0.217) | 0.867<br>(0.217) | 0.867<br>(0.139) | 0.900<br>(0.149) | 0,871 | 0,848 | 0,85 | 0,856 | 0.867<br>(0.217) | 0.833<br>(0.236) | 0.600<br>(0.091) | 0.867<br>(0.217) | 0,886 | 0,833 | 0,845 | 0,83 |
|  | Se | 0.800<br>(0.274) | 0.800<br>(0.274) | 0.700<br>(0.274) | 0.800<br>(0.274) | 0,75 | 0,5 | 0,5 | 0 | 0.800<br>(0.274) | 0.800<br>(0.274) | 0.700<br>(0.274) | 0.800<br>(0.274) | 0,708 | 0,5 | 0,583 | 0 |
|  | Sp | 0.733<br>(0.149) | 1.000<br>(0.000) | 0.600<br>(0.279) | 0.800<br>(0.298) | 0,818 | 0,909 | 0,909 | 1 | 0.600<br>(0.279) | 0.667<br>(0.236) | 0.333<br>(0.236) | 0.867<br>(0.183) | 0,909 | 0,818 | 1 | 1 |

HC: healthy controls; PD: Parkinson's disease; PSP: Progressive supranuclear palsy; MSA-C: cerebellar variant of multiple system atrophy; MSA-P: parkinsonian variant of multiple system atrophy; DTI: diffusion tensor imaging; BA: balanced accuracy; Se: sensitivity; Sp: specificity; SVM: support-vector machine

**Table S4:**
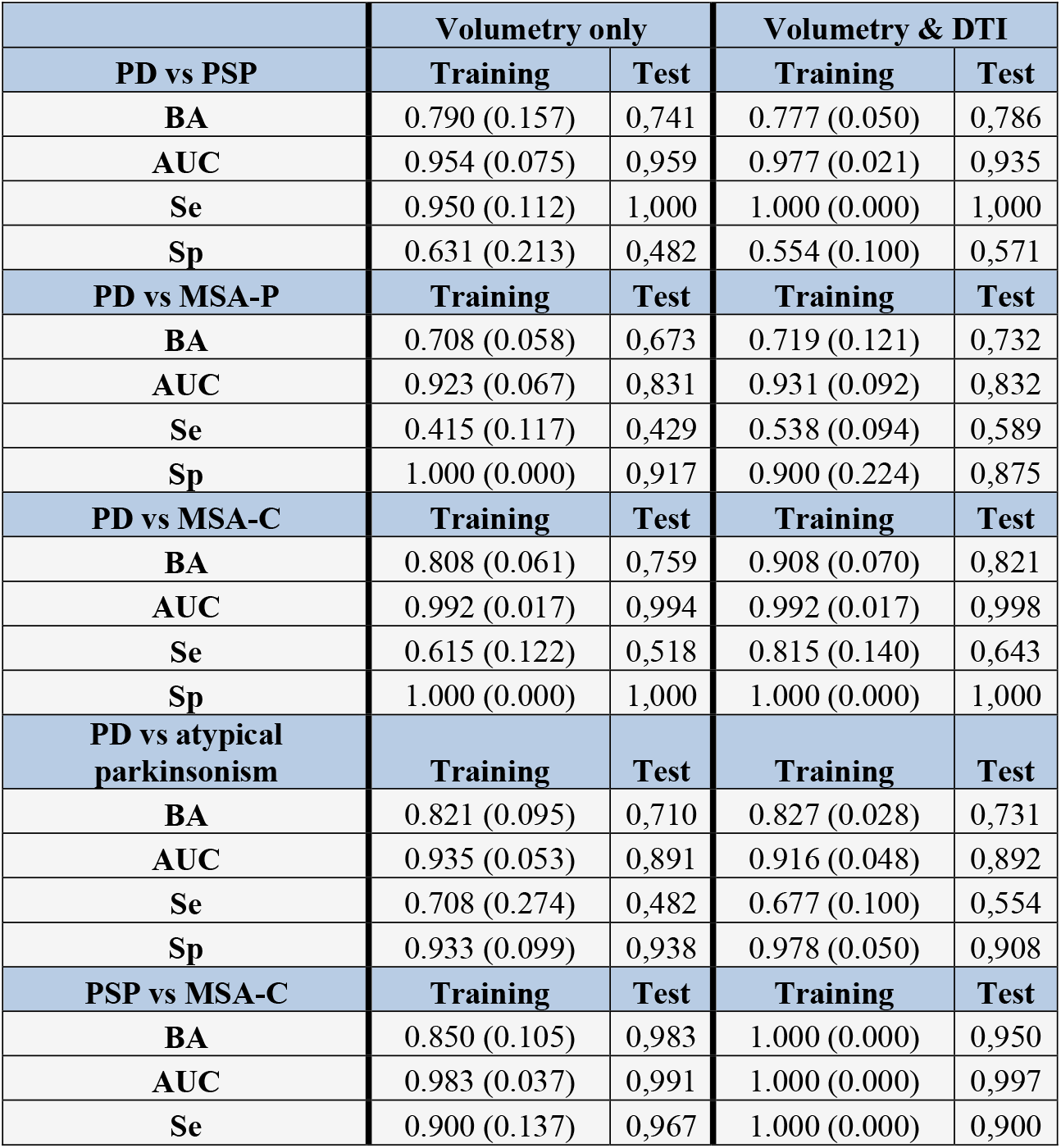
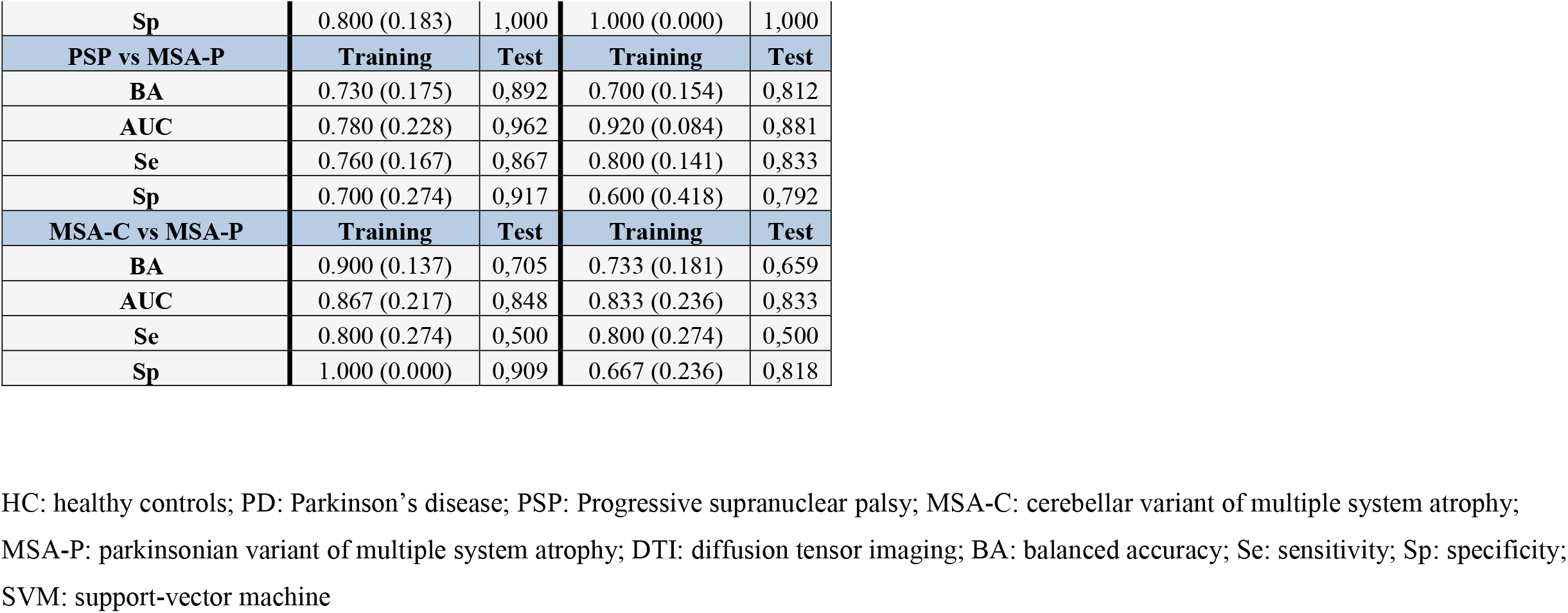
Performances of SVM classification in the training cohort and in the subset of the replication cohort after normalization using HCs for volumetric data; then for the combination of volumetric and DTI metrics.

|  | Volumetry only |  | Volumetry & DTI |  |
| --- | --- | --- | --- | --- |
| PD vs PSP | Training | Test | Training | Test |
| BA | 0.790 (0.157) | 0,741 | 0.777 (0.050) | 0,786 |
| AUC | 0.954 (0.075) | 0,959 | 0.977 (0.021) | 0,935 |
| Se | 0.950 (0.112) | 1,000 | 1.000 (0.000) | 1,000 |
| Sp | 0.631 (0.213) | 0,482 | 0.554 (0.100) | 0,571 |
| PD vs MSA-P | Training | Test | Training | Test |
| BA | 0.708 (0.058) | 0,673 | 0.719 (0.121) | 0,732 |
| AUC | 0.923 (0.067) | 0,831 | 0.931 (0.092) | 0,832 |
| Se | 0.415 (0.117) | 0,429 | 0.538 (0.094) | 0,589 |
| Sp | 1.000 (0.000) | 0,917 | 0.900 (0.224) | 0,875 |
| PD vs MSA-C | Training | Test | Training | Test |
| BA | 0.808 (0.061) | 0,759 | 0.908 (0.070) | 0,821 |
| AUC | 0.992 (0.017) | 0,994 | 0.992 (0.017) | 0,998 |
| Se | 0.615 (0.122) | 0,518 | 0.815 (0.140) | 0,643 |
| Sp | 1.000 (0.000) | 1,000 | 1.000 (0.000) | 1,000 |
| PD vs atypical parkinsonism | Training | Test | Training | Test |
| BA | 0.821 (0.095) | 0,710 | 0.827 (0.028) | 0,731 |
| AUC | 0.935 (0.053) | 0,891 | 0.916 (0.048) | 0,892 |
| Se | 0.708 (0.274) | 0,482 | 0.677 (0.100) | 0,554 |
| Sp | 0.933 (0.099) | 0,938 | 0.978 (0.050) | 0,908 |
| PSP vs MSA-C | Training | Test | Training | Test |
| BA | 0.850 (0.105) | 0,983 | 1.000 (0.000) | 0,950 |
| AUC | 0.983 (0.037) | 0,991 | 1.000 (0.000) | 0,997 |
| Se | 0.900 (0.137) | 0,967 | 1.000 (0.000) | 0,900 |
| **Sp** | 0.800 (0.183) | 1,000 | 1.000 (0.000) | 1,000 |
| **PSP vs MSA-P** | **Training** | **Test** | **Training** | **Test** |
| **BA** | 0.730 (0.175) | 0,892 | 0.700 (0.154) | 0,812 |
| **AUC** | 0.780 (0.228) | 0,962 | 0.920 (0.084) | 0,881 |
| **Se** | 0.760 (0.167) | 0,867 | 0.800 (0.141) | 0,833 |
| **Sp** | 0.700 (0.274) | 0,917 | 0.600 (0.418) | 0,792 |
| **MSA-C vs MSA-P** | **Training** | **Test** | **Training** | **Test** |
| **BA** | 0.900 (0.137) | 0,705 | 0.733 (0.181) | 0,659 |
| **AUC** | 0.867 (0.217) | 0,848 | 0.833 (0.236) | 0,833 |
| **Se** | 0.800 (0.274) | 0,500 | 0.800 (0.274) | 0,500 |
| **Sp** | 1.000 (0.000) | 0,909 | 0.667 (0.236) | 0,818 |

HC: healthy controls; PD: Parkinson's disease; PSP: Progressive supranuclear palsy; MSA-C: cerebellar variant of multiple system atrophy; MSA-P: parkinsonian variant of multiple system atrophy; DTI: diffusion tensor imaging; BA: balanced accuracy; Se: sensitivity; Sp: specificity; SVM: support-vector machine

## Abbreviations and acronyms

(AUC): Area under the ROC curve
(ADC): Apparent diffusion coefficient
(AD): Axial diffusivity
(BA): Balanced accuracy
(DTI): Diffusion tensor imaging
(FA): Fractional anisotropy
(V4): Fourth ventricle
(MD): Mean diffusivity
(MRI): Magnetic resonance imaging
(MSA-C): Cerebellar variant of multiple system atrophy
(MSA-P): Parkinsonian variant of multiple system atrophy
(PD): Parkinson’s disease
(PSP): Progressive supranuclear palsy
(RD): Radial diffusivity
(SVM): Support vector machine
(SCP): Superior cerebellar peduncles
(V3): Third ventricle
(UPDRS): Unified Parkinson’s Disease Rating Scale

---

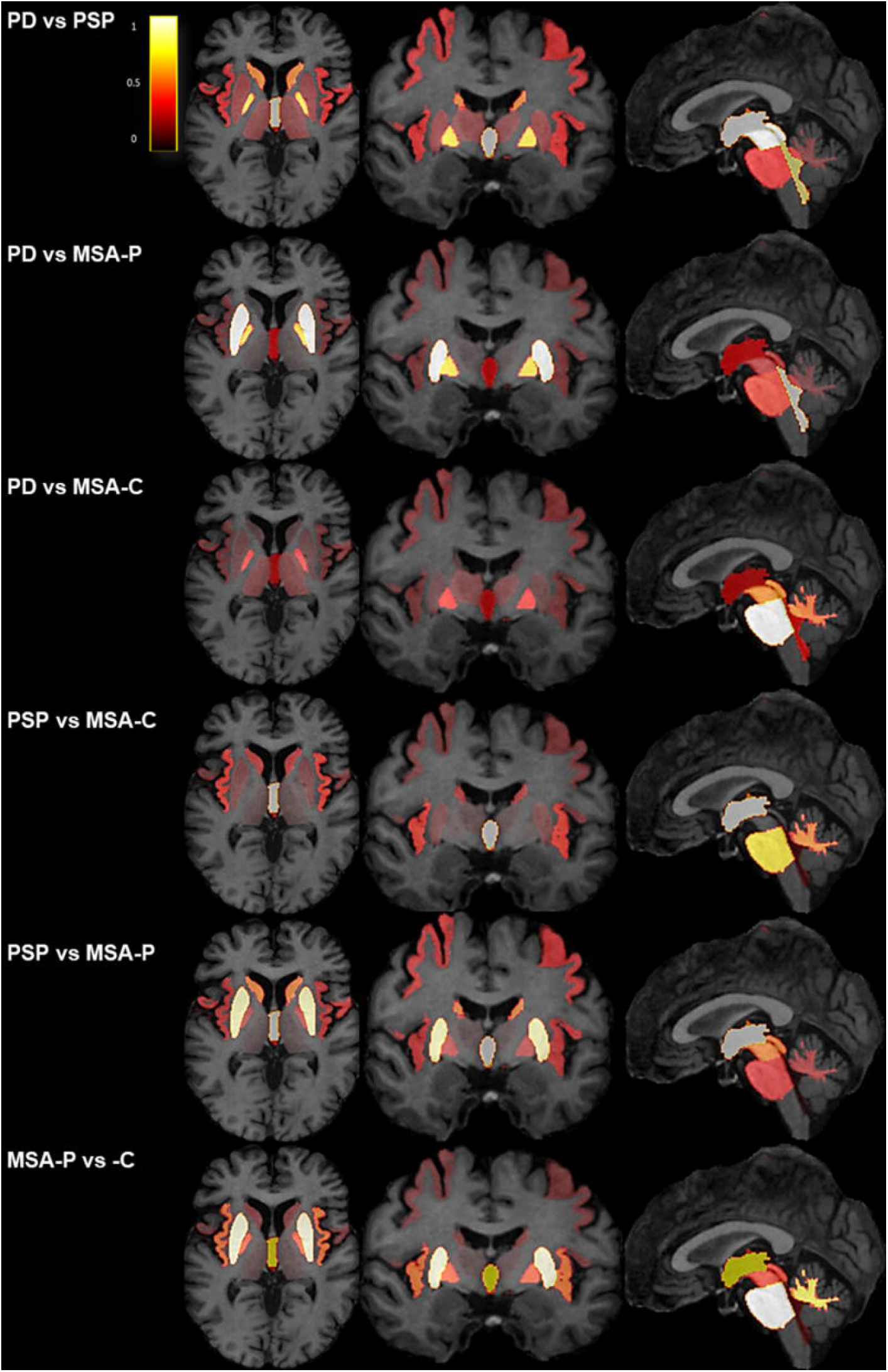

